# Innovative COVID-19 Point-of-Care Diagnostics Suitable for Tuberculosis Diagnosis: A Scoping Review

**DOI:** 10.1101/2024.06.13.24308880

**Authors:** Lydia Holtgrewe, Sonal Jain, Ralitza Dekova, Tobias Broger, Chris Isaacs, Payam Nahid, Adithya Cattamanchi, Claudia M. Denkinger, Seda Yerlikaya

## Abstract

**Introduction:** Rapid and accurate point-of-care (POC) tuberculosis (TB) diagnostics are a key priority to close the TB diagnostic gap of 3.1 million people without a diagnosis. Leveraging the recent surge in COVID-19 diagnostic innovation, we explored the potential adaptation of commercially available SARS-CoV-2 tests for TB diagnosis, aligning with World Health Organization (WHO) target product profiles (TPPs).

**Methods:** A scoping review was conducted following PRISMA-ScR guidelines to systematically map commercially available POC molecular and antigen SARS-CoV-2 diagnostic tests potentially meeting the TPPs for TB diagnostic tests for peripheral settings. Data were gathered from PubMed/MEDLINE, bioRxiv, and medRxiv, along with publicly accessible in vitro diagnostic test databases, and developer websites, up to November 23, 2022. Data on developer and test attributes, operational characteristics, pricing, and clinical performance were charted using standardized data extraction forms. Each identified test was evaluated using a standardized scorecard. A narrative synthesis of the charted data is presented.

**Results:** Our database search yielded 2,003 studies, from which 408 were considered eligible. Among these, we identified 58 commercialized diagnostic devices, including 17 near-POC antigen tests, one POC molecular test, 29 near-POC molecular tests, and 11 low-complexity molecular tests. We summarized the detailed characteristics, regulatory status, and clinical performance data of these tests. The LumiraDx (Roche, Switzerland) emerged as the highest-scoring near-POC antigen platform, while Visby (Visby, USA) was the highest-performing near-POC molecular platform. The Lucira Check-It (Pfizer, USA) was noted as the sole POC molecular test. The Idylla^TM^ (Biocartis, Switzerland) was identified as the leading low-complexity molecular test.

**Discussion:** We highlight a diverse landscape of commercially available diagnostic tests suitable for potential adaptation to TB POC testing. This work aims to bolster global TB initiatives by fostering stakeholder collaboration, leveraging COVID-19 diagnostic technologies for TB diagnosis, and uncovering new commercial avenues to tackle longstanding challenges in TB diagnosis.

## 1 INTRODUCTION

### 1.1 RATIONALE

While healthcare systems are slowly recovering from the disruptive impacts of the COVID- 19 pandemic, tuberculosis (TB) remains the world’s leading infectious killer. It accounted for 1.6 million new cases and 1.3 million deaths in 2022 alone[1]. Despite a modest 8.7% reduction in TB incidence between 2015 and 2022, the global community remains far away from achieving the 50% reduction target set by the World Health Organization (WHO) End TB Strategy for 2025[1]. To reach this milestone and to curb the community transmission of TB, effective diagnosis and treatment of people with TB is essential [2]. Closing the many gaps in the TB care cascade necessitates improvements in access to TB testing, optimized utilization of diagnostic tests, and strengthening the linkage to TB treatment[3]. Bridging the diagnostic gap requires rapid, accurate, and affordable diagnostic tests suitable for use at point-of-care (POC).

In an effort to guide developers towards fit-for-purpose TB diagnostics, the WHO defined high priority target product profiles (TPP) in 2014[4]. Currently, a revision of these TPPs is underway, with a preliminary draft having been shared in August 2023 as part of a public consultation[5]. Current WHO-recommended rapid diagnostic tests (WRDs) fall short of the minimal TPP requirements for a TB diagnostic for peripheral settings, either due to a reliance on sputum specimens, inadequate clinical performance, cost, and/or limited operational suitability[4, 6, 7]. As a result, as of 2022, only 47% of notified TB cases worldwide were diagnosed using WRDs[8–10]. Fit-for-purpose POC TB diagnostic tests that meet TPP criteria are needed to achieve the WHO’s goal of attaining 100% global coverage of WRDs[10].

Leveraging the rapid advances in diagnostic technology as a result of the COVID-19 pandemic offers the potential to rapidly develop better solutions for TB diagnosis. Through increased funding and collaborative initiatives, such as the Access to COVID-19 Tools (ACT) Accelerator or RADx, the pandemic created growth opportunities for key market players and fueled an unprecedented momentum in the research and development (R&D) of novel diagnostics[11]. This resulted in the development of a diverse array of diagnostic products for remote and at-home testing[12]. As the size of the global COVID-19 diagnostics market is declining [11], developers are looking for new avenues to apply their innovations. TB can emerge as a practical choice for these developers given the substantial disease burden, supportive government initiatives, and in-kind funding to support validation through established research networks.

### 1.2 OBJECTIVES

This scoping review was conducted to systematically map the commercially available molecular and antigen SARS-CoV-2 diagnostic tests with the potential of meeting the 2014 TPP for new POC TB diagnostics [4]. This compilation of commercialized diagnostic tools seeks to identify promising innovations to facilitate interactions between device and assay developers with other key stakeholders, leveraging the momentum in diagnostic innovation spurred by COVID-19 to address gaps in TB diagnostics.

## 2 METHODS

This is a scoping review of the scientific literature, SARS-CoV-2 test databases, and information made available by developers. It follows the PRISMA Extension for Scoping Reviews (PRISMA-ScR) guidelines (see Table S1), and the methodological framework developed by Levac et al[13, 14].

### 2.1 PROTOCOL AND REGISTRATION

We previously published the protocol for this scoping review[15]. Because of the vast amount of identified literature, we split the work into two, with this publication focusing exclusively on commercialized diagnostics and a publication (in preparation) pertaining to the tests that are either in the pre-commercialization stage or are still in development.

### 2.2 DEFINITIONS AND ELIGIBILITY CRITERIA

The definitions and eligibility criteria for diagnostic devices used in this work are defined in the protocol[15]. We added the following sub-categories for peripheral in vitro diagnostic (IVD) tests:

● POC tests: Diagnostic tests performed at or near the site of patient care. These tests are designed to be instrument-free, disposable, and do not require specific infrastructure, such as access to mains electricity, laboratory equipment, or a cold chain. They can be placed in healthcare settings without laboratory infrastructure and do not require any special skills to administered. Example: Alere Determine TB LAM Ag Test (Abbott, IL, USA).
● Near-POC tests: Diagnostics tests that are instrument-based and only require basic infrastructure, such as mains electricity for recharging batteries or operating instruments. These test can be used in healthcare settings without laboratories and can be performed by healthcare workers with basic technical skills, such as simple pipetting and sample transfer that do not require precise timing or volumes. Ideally, these tests come with transfer pipettes with pre-set volumes provided by the manufacturer to avoid the need for laboratory skills. Example: GeneXpert Edge (Cepheid, CA, USA).
● Low-complexity tests: Instrument-based tests intended for use in healthcare settings with basic laboratory infrastructure and access to mains electricity. These tests require basic technical skills and laboratory equipment, including pipettes, vortex mixers, heating devices, freezers, and separate test tubes. Examples: Truenat (Molbio Diagnostics, India), GeneXpert 6-/10-color platforms (Cepheid, CA, USA).

We made the following modification to the eligibility criteria of POC molecular and antigen tests used for SARS-CoV-2 detection in the scoping review protocol[15]:

- ● Minimal biosafety requirements (e.g., personal protective equipment (PPE), good ventilation, and a biohazard bag for waste disposal): because only very few devices reported data on this parameter, we decided to omit it from the data collection.

### 2.3 INFORMATION SOURCES

As specified in the protocol, we initially searched for relevant peer-reviewed literature in PubMed/MEDLINE and for pre-prints in bioRxiv and medRxiv[15]. We then searched additional IVD databases and developer websites to obtain supplementary information on each diagnostic test identified through the above-mentioned databases. We updated the website links to the following IVD databases:

● U.S. Food and Drug Administration (FDA) Tables of In Vitro Diagnostics Emergency Use Authorizations: https://www.fda.gov/medical-devices/covid-19-emergency-use-authorizations-medical-devices/in-vitro-diagnostics-euas-molecular-diagnostic-tests-sars-cov-2.
● National Institutes of Health (NIH) Rapid Acceleration of Diagnostics (RADx®): https://www.nibib.nih.gov/covid-19/radx-tech-program/authorized-tests.

The NMPA - China Medical Products Administration Database (https://www.nmpa.gov.cn/datasearch/en/search-result-en.html?nmpaItem<u>=</u> <u>82808081889a0b5601889a251e33005c</u>) and CDSCO - Government of India, Central Drugs Standard Control Organization (https://cdsco.gov.in/opencms/opencms/en/Medical-Device-Diagnostics/InVitro-Diagnostics/) had limited search function and language barriers and the majority of devices could not be identified. The EUDAMED (https://ec.europa.eu/tools/eudamed/#/screen/search-device) was not accessible for data search at the time of data collection. [16].

### 2.4 SEARCH STRATEGY

The full search term for PubMed/MEDLINE is shown in Table 1 in the protocol[15]. It was adapted as necessary for the other databases bioRxiv and medRxiv using the medrxivr package in R (version 4.0.5; R Foundation for Statistical Computing) to overcome the limitations of the search functionality of these websites and allow for reproducibility. The analytical R code can be found in the Supplementary Methods, Section 1. We did not impose any restrictions on the publication date or language of relevant manuscripts.

**Table 1.**
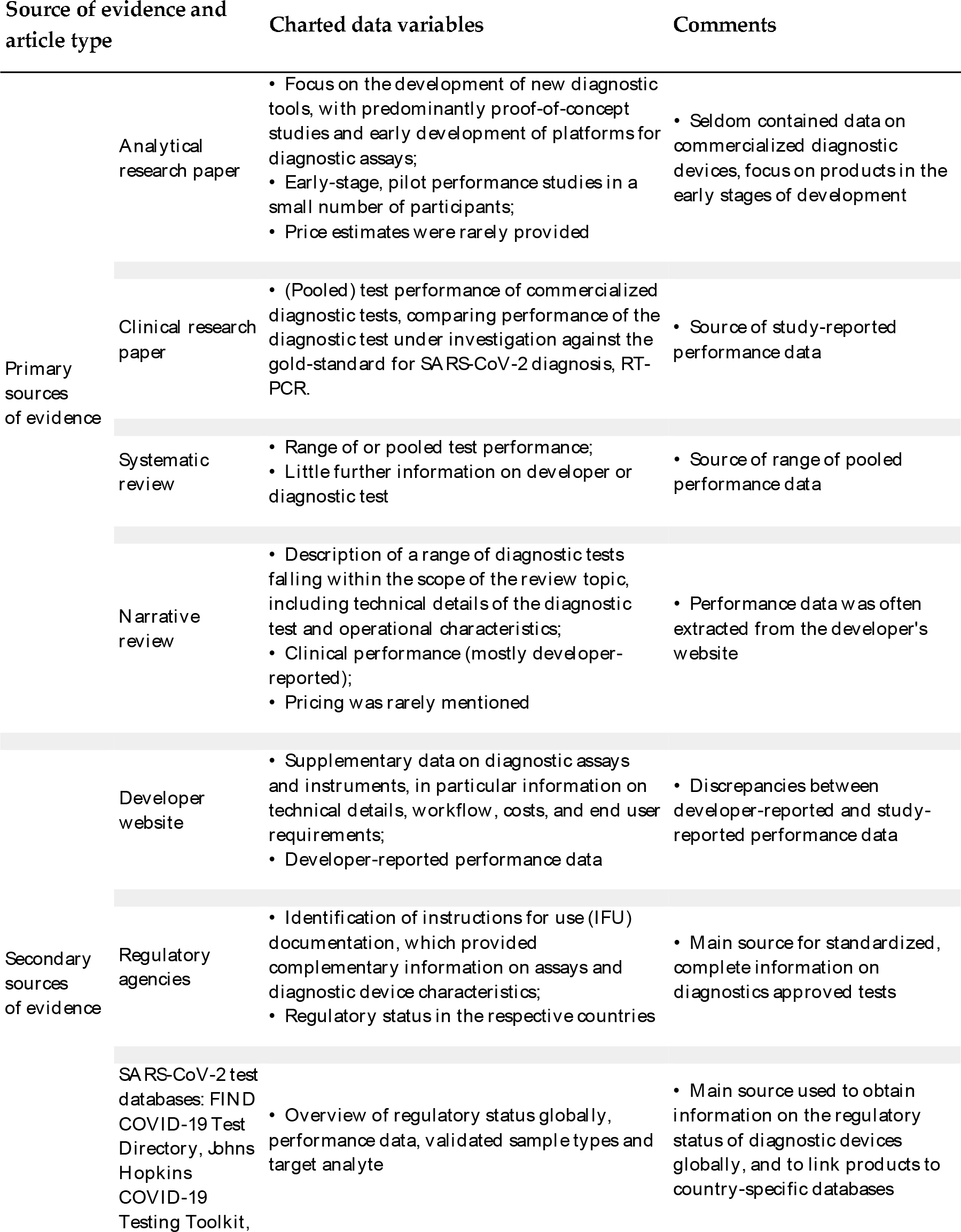
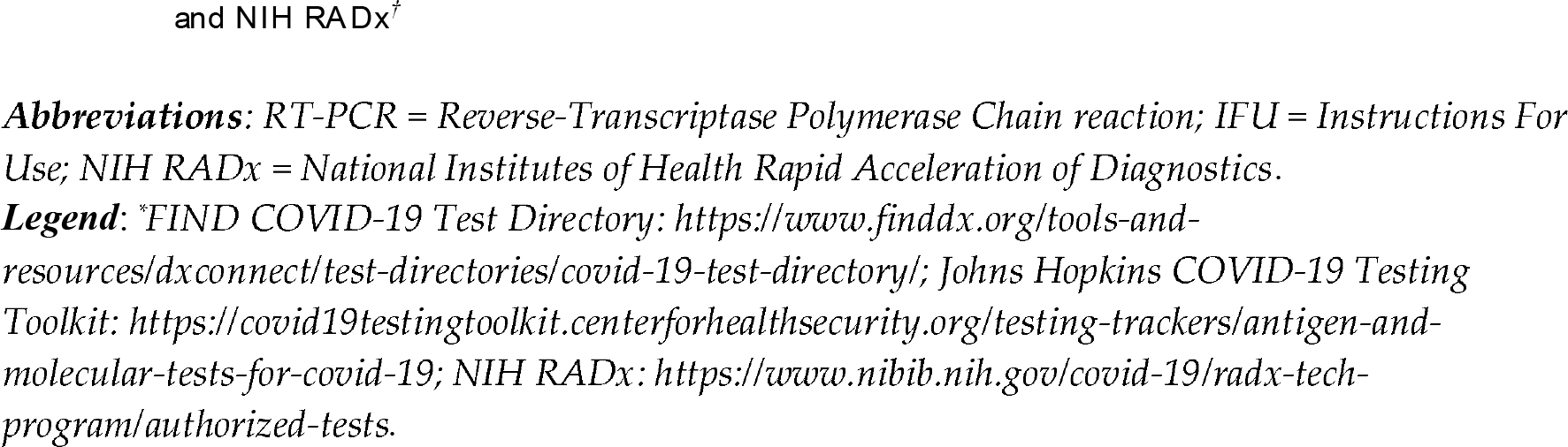
Characteristics of included sources of evidence and charted data variables.

### 2.5 SELECTION OF SOURCES OF EVIDENCE

Retrieved articles were collated using Covidence systematic review software (Veritas Health Innovation, Melbourne, Australia; available at www.covidence.com) and duplicates were automatically removed[17]. The same software was used for screening. Two reviewers (S.Y., L.H.) independently screened titles and abstracts against the eligibility criteria. Then, full-text screening was performed by the same reviewers. Any discrepancies were resolved through consensus.

### 2.6 DATA CHARTING PROCESS

We used two Google forms for data charting. The forms were developed by one reviewer (S.Y.) and revised in an iterative process by both reviewers (S.Y., L.H.) during initial data charting. One reviewer (L.H.) charted information on study design, test characteristics, and clinical performance from eligible studies using the first standardized form (see Table S2). For studies that mentioned more than one test, multiple records were charted (one record per test). The same reviewer (L.H.) used the second standardized form to collect additional information on tests identified during the first charting step, drawing on additional sources, such as the developer’s website, and the regulatory databases listed above (see Table S3). Due to a high number of extracted publications, only the results tables, and not the primary data, were cross-checked by a second reviewer (S.J. and R.D.). Data charted from various sources were collated on separate excel sheets for each diagnostic test.

### 2.7 VARIABLES

We abstracted data on test description, operation characteristics, pricing, performance, and commercialization status, as listed in Table 2 in the study protocol[15]. Variables are defined in the data charting forms.

**Table 2.**
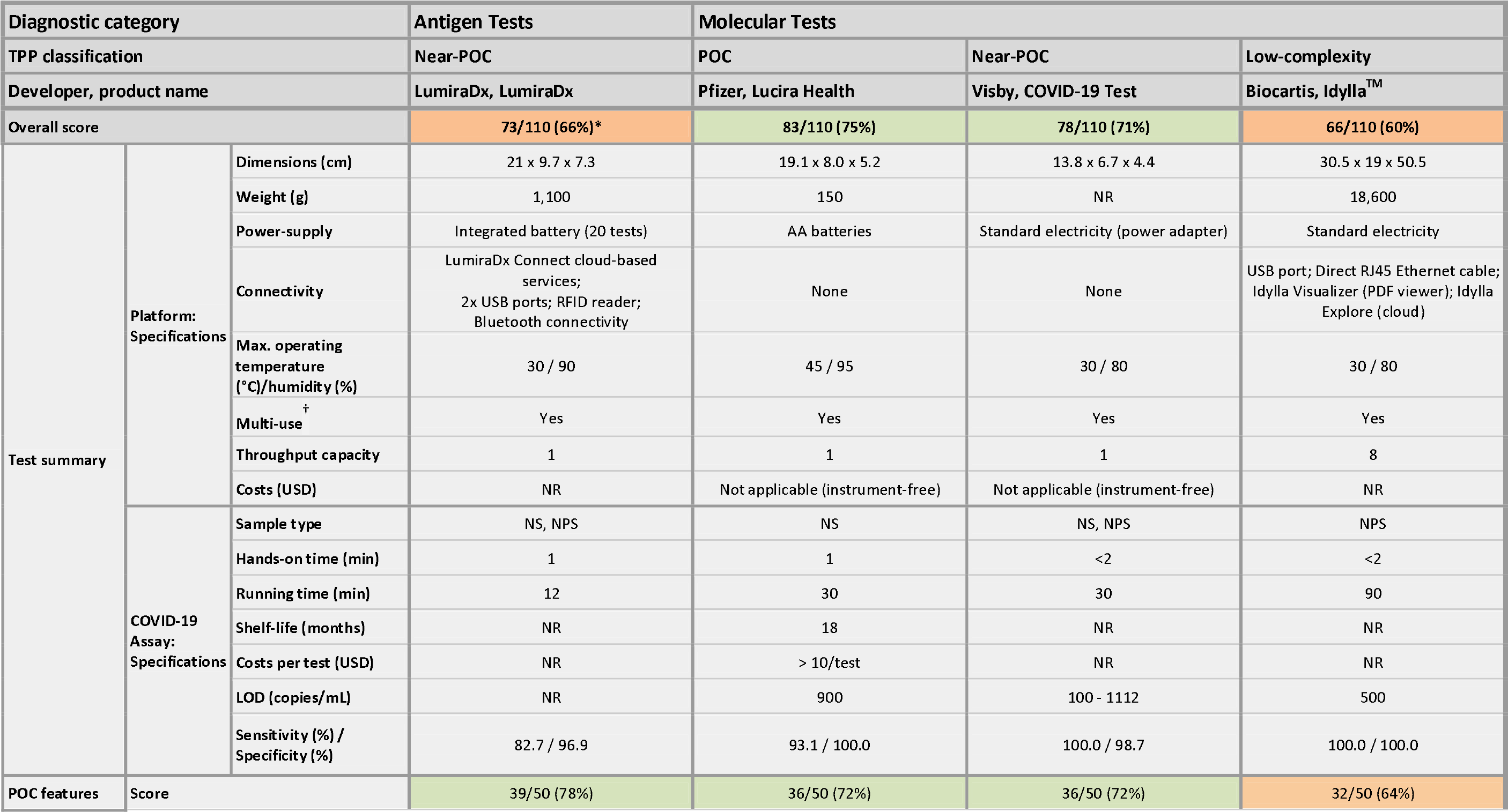
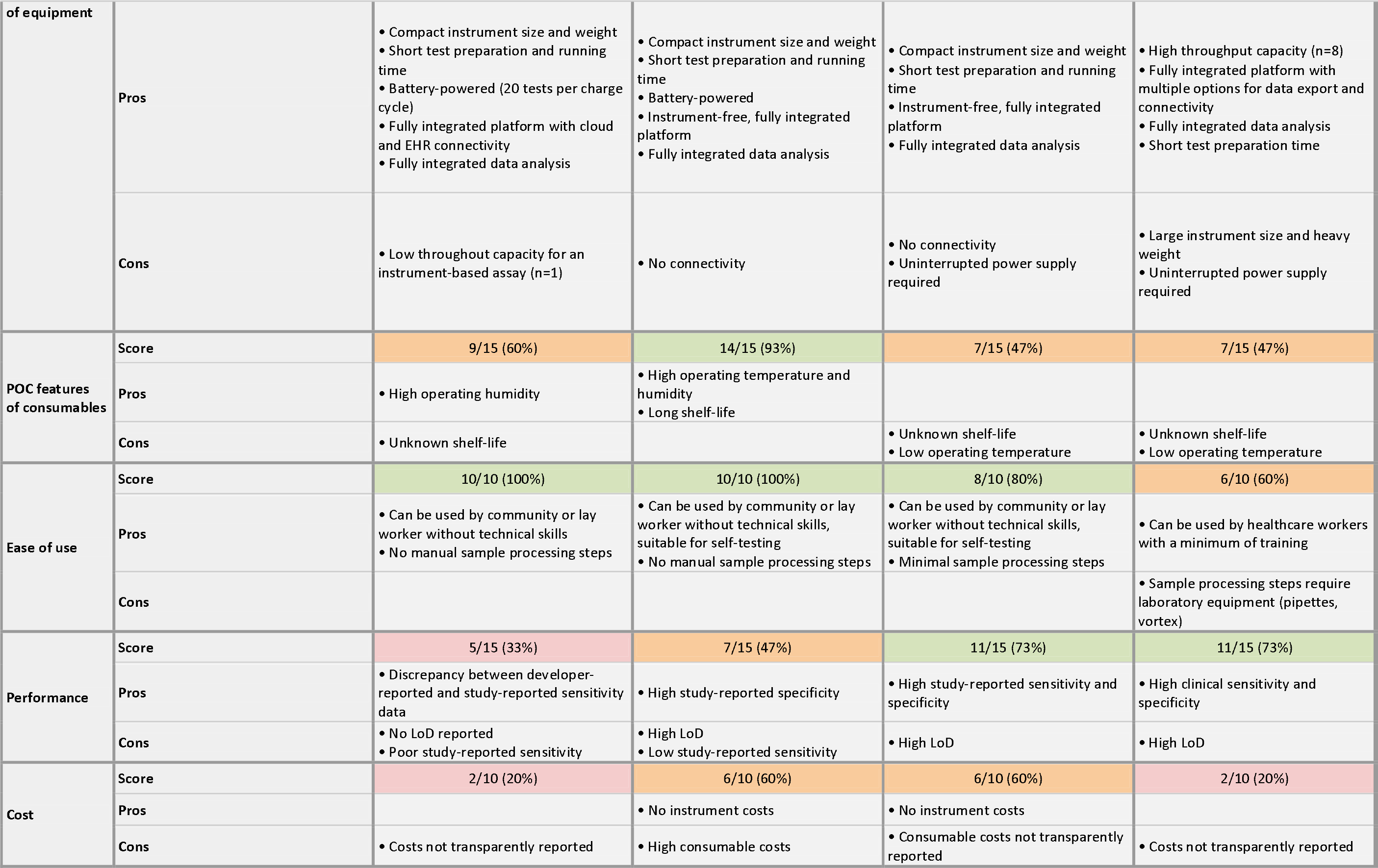
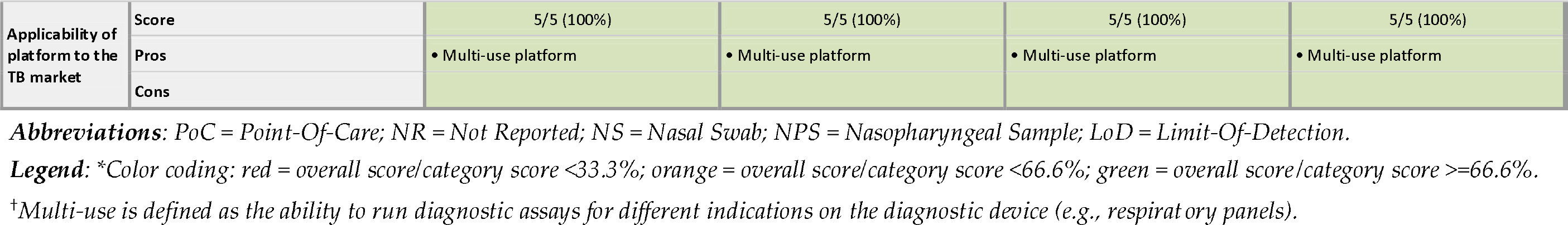
Characteristics of highest-scoring diagnostic devices stratified by test category.

| Diagnostic category |  |  | Antigen Tests | Molecular Tests |  |  |
| --- | --- | --- | --- | --- | --- | --- |
| TPP classification |  |  | Near-POC | POC | Near-POC | Low-complexity |
| Developer, product name |  |  | LumiraDx, LumiraDx | Pfizer, Lucira Health | Visby, COVID-19 Test | Biocartis, Idylla™ |
| Overall score |  |  | 73/110 (66%)* | 83/110 (75%) | 78/110 (71%) | 66/110 (60%) |
| Test summary | Platform: Specifications | Dimensions (cm) | 21 x 9.7 x 7.3 | 19.1 x 8.0 x 5.2 | 13.8 x 6.7 x 4.4 | 30.5 x 19 x 50.5 |
|  |  | Weight (g) | 1,100 | 150 | NR | 18,600 |
|  |  | Power-supply | Integrated battery (20 tests) | AA batteries | Standard electricity (power adapter) | Standard electricity |
|  |  | Connectivity | LumiraDx Connect cloud-based services;<br>2x USB ports; RFID reader;<br>Bluetooth connectivity | None | None | USB port; Direct RJ45 Ethernet cable;<br>Idylla Visualizer (PDF viewer); Idylla Explore (cloud) |
|  |  | Max. operating temperature (°C)/humidity (%) | 30 / 90 | 45 / 95 | 30 / 80 | 30 / 80 |
|  |  | Multi-use<sup>†</sup> | Yes | Yes | Yes | Yes |
|  |  | Throughput capacity | 1 | 1 | 1 | 8 |
|  |  | Costs (USD) | NR | Not applicable (instrument-free) | Not applicable (instrument-free) | NR |
|  | COVID-19 Assay: Specifications | Sample type | NS, NPS | NS | NS, NPS | NPS |
|  |  | Hands-on time (min) | 1 | 1 | <2 | <2 |
|  |  | Running time (min) | 12 | 30 | 30 | 90 |
|  |  | Shelf-life (months) | NR | 18 | NR | NR |
|  |  | Costs per test (USD) | NR | > 10/test | NR | NR |
|  |  | LOD (copies/mL) | NR | 900 | 100 - 1112 | 500 |
|  |  | Sensitivity (%) / Specificity (%) | 82.7 / 96.9 | 93.1 / 100.0 | 100.0 / 98.7 | 100.0 / 100.0 |
| POC features | Score |  | 39/50 (78%) | 36/50 (72%) | 36/50 (72%) | 32/50 (64%) |

|  |  |  |  |  |  |
| --- | --- | --- | --- | --- | --- |
| of equipment | Pros | • Compact instrument size and weight<br>• Short test preparation and running time<br>• Battery-powered (20 tests per charge cycle)<br>• Fully integrated platform with cloud and EHR connectivity<br>• Fully integrated data analysis | • Compact instrument size and weight<br>• Short test preparation and running time<br>• Battery-powered<br>• Instrument-free, fully integrated platform<br>• Fully integrated data analysis | • Compact instrument size and weight<br>• Short test preparation and running time<br>• Instrument-free, fully integrated platform<br>• Fully integrated data analysis | • High throughput capacity (n=8)<br>• Fully integrated platform with multiple options for data export and connectivity<br>• Fully integrated data analysis<br>• Short test preparation time |
|  | Cons | • Low throughput capacity for an instrument-based assay (n=1) | • No connectivity | • No connectivity<br>• Uninterrupted power supply required | • Large instrument size and heavy weight<br>• Uninterrupted power supply required |
| POC features of consumables | Score | 9/15 (60%) | 14/15 (93%) | 7/15 (47%) | 7/15 (47%) |
|  | Pros | • High operating humidity | • High operating temperature and humidity<br>• Long shelf-life |  |  |
|  | Cons | • Unknown shelf-life |  | • Unknown shelf-life<br>• Low operating temperature | • Unknown shelf-life<br>• Low operating temperature |
| Ease of use | Score | 10/10 (100%) | 10/10 (100%) | 8/10 (80%) | 6/10 (60%) |
|  | Pros | • Can be used by community or lay worker without technical skills<br>• No manual sample processing steps | • Can be used by community or lay worker without technical skills, suitable for self-testing<br>• No manual sample processing steps | • Can be used by community or lay worker without technical skills, suitable for self-testing<br>• Minimal sample processing steps | • Can be used by healthcare workers with a minimum of training |
|  | Cons |  |  |  | • Sample processing steps require laboratory equipment (pipettes, vortex) |
| Performance | Score | 5/15 (33%) | 7/15 (47%) | 11/15 (73%) | 11/15 (73%) |
|  | Pros | • Discrepancy between developer-reported and study-reported sensitivity data | • High study-reported specificity | • High study-reported sensitivity and specificity | • High clinical sensitivity and specificity |
|  | Cons | • No LoD reported<br>• Poor study-reported sensitivity | • High LoD<br>• Low study-reported sensitivity | • High LoD | • High LoD |
| Cost | Score | 2/10 (20%) | 6/10 (60%) | 6/10 (60%) | 2/10 (20%) |
|  | Pros |  | • No instrument costs | • No instrument costs |  |
|  | Cons | • Costs not transparently reported | • High consumable costs | • Consumable costs not transparently reported | • Costs not transparently reported |
| Applicability of platform to the TB market | Score | 5/5 (100%) | 5/5 (100%) | 5/5 (100%) | 5/5 (100%) |
|  | Pros | • Multi-use platform | • Multi-use platform | • Multi-use platform | • Multi-use platform |
|  | Cons |  |  |  |  |

**Abbreviations:** PoC = Point-Of-Care; NR = Not Reported; NS = Nasal Swab; NPS = Nasopharyngeal Sample; LoD = Limit-Of-Detection.
**Legend:** \*Color coding: red = overall score/category score <33.3%; orange = overall score/category score <66.6%; green = overall score/category score >=66.6%.
<sup>†</sup>Multi-use is defined as the ability to run diagnostic assays for different indications on the diagnostic device (e.g., respiratory panels).

### 2.8 SYNTHESIS OF RESULTS

In accordance with current standards for scoping reviews and evidence mapping, a narrative synthesis of information is provided in the text and tables to summarize and explain major aspects of the included diagnostic tests, such as information on the developer, test characteristics, and clinical performance data stratified by technology type (antigen and molecular) and test classification (low-complexity, near-POC, and POC).

We modified Lehe et al.’s standardized scorecard, created for the objective evaluation of operational characteristics of POC diagnostic devices, to align with the diagnostic requirements of the WHO’s TPP in order to reduce subjectivity in the assessment of the included SARS-CoV-2 diagnostic devices [18]. Diagnostic devices received an overall score of a minimum of zero to a maximum of 110 points based on 22 scoring criteria. Scoring criteria were classified into seven scoring categories. Each scoring criterion was assigned one to five points, with one being the lowest, and five being the highest possible score. When information was missing, the lowest score of one point was given. The adapted scoring framework, including definitions of all scoring criteria, is shown in the Table S4. Each diagnostic device was independently scored by two reviewers (L.H., S.J.) and scoring conflicts were resolved through consensus and discussion with a third reviewer (S.Y.). We present device characteristics and performance of the highest-scoring diagnostic tests in each of the six categories in tables, figures, and text.

## 3 RESULTS

### 3.1 SELECTION OF SOURCES OF EVIDENCE

Database search yielded 2,003 results that were imported into Covidence for screening. After 248 duplicates were automatically removed by Covidence, 1754 studies underwent title/abstract screening, out of which 874 were considered eligible for full-text screening. Among these, the most common reasons for exclusion were inclusion of assays and instruments that were ineligible for use in peripheral settings (n = 234), no mention of specific tests (n = 127) and reporting on conventional lateral flow assays without reading devices or enhanced detection technologies (n = 56), as shown in Fig. 1. From the 408 studies considered eligible for inclusion in this review, 58 commercialized diagnostic devices were identified using the primary sources of evidence.

**Fig. 1.**
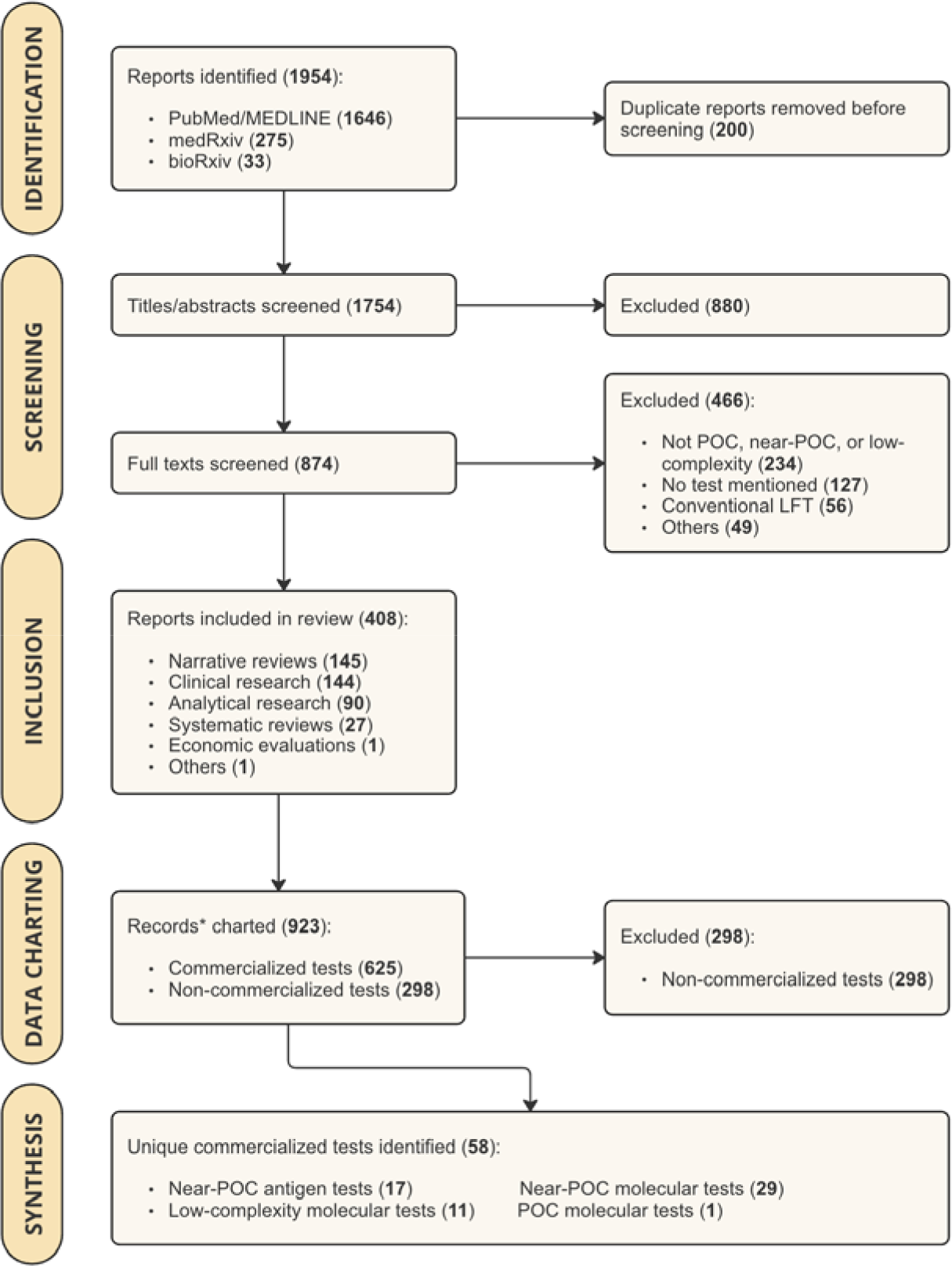
PRISMA Flow Chart, showing the results of study search and screening procedures. Abbreviations: PoC = Point-Of-Care; LFT = Lateral Flow *Test; PCR = Polymerase Chain Reaction.* ***Legend****: *For studies that mentioned more than one tests, multiple records were charted (one record per test)*.

### 3.2 CHARACTERISTICS OF INDIVIDUAL SOURCES OF EVIDENCE

The data charted from included evidence sources is described in Table 1, along with comments on the source characteristics.

### 3.3 SYNTHESIS OF RESULTS

Out of 58 commercialized POC diagnostic tests for SARS-CoV-2, we identified 17 near-POC antigen tests, one POC molecular test, 29 near-POC molecular tests, and 11 low-complexity molecular tests. By definition, there were no POC antigen tests in the review because we excluded conventional instrument-free LFTs. The 55 manufacturers of included diagnostic tests are displayed in Fig. 2. Developer and product characteristics, regulatory status, and clinical performance data of all included diagnostic tests are shown in Table S5-S8. Table 2 displays platform and assay characteristics, clinical performance, and end-user requirements of the single highest-scoring diagnostic test within each diagnostic category. For each of the three highest-scoring diagnostic tests within each diagnostic category, Fig. 3 shows the scores across all seven categories.

**Fig. 2.**
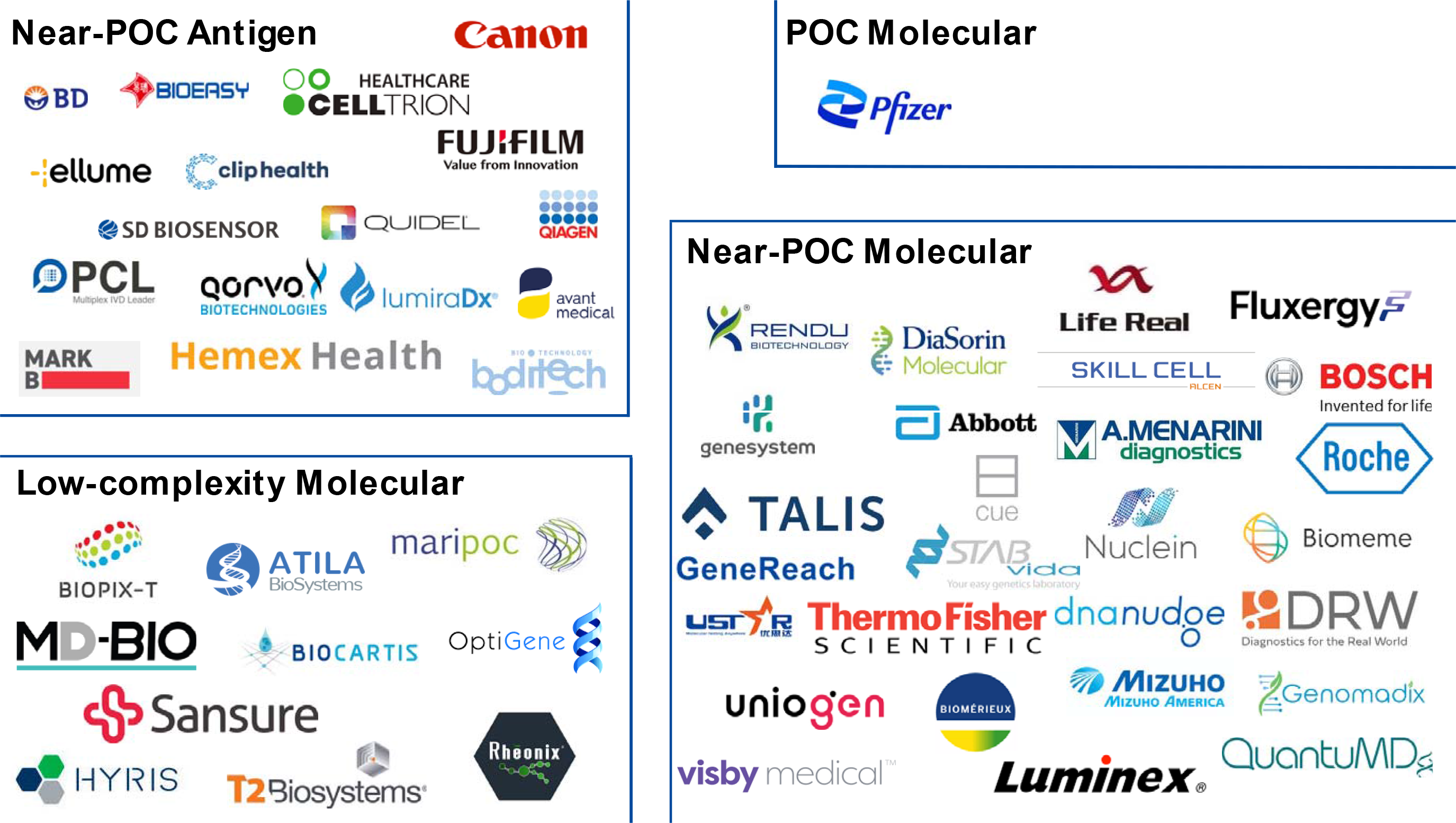
Logo chart, displaying the 55 manufacturers of all 58 included diagnostic tests. Abbreviations: PoC = Point-Of-Care.

**Fig 3.**
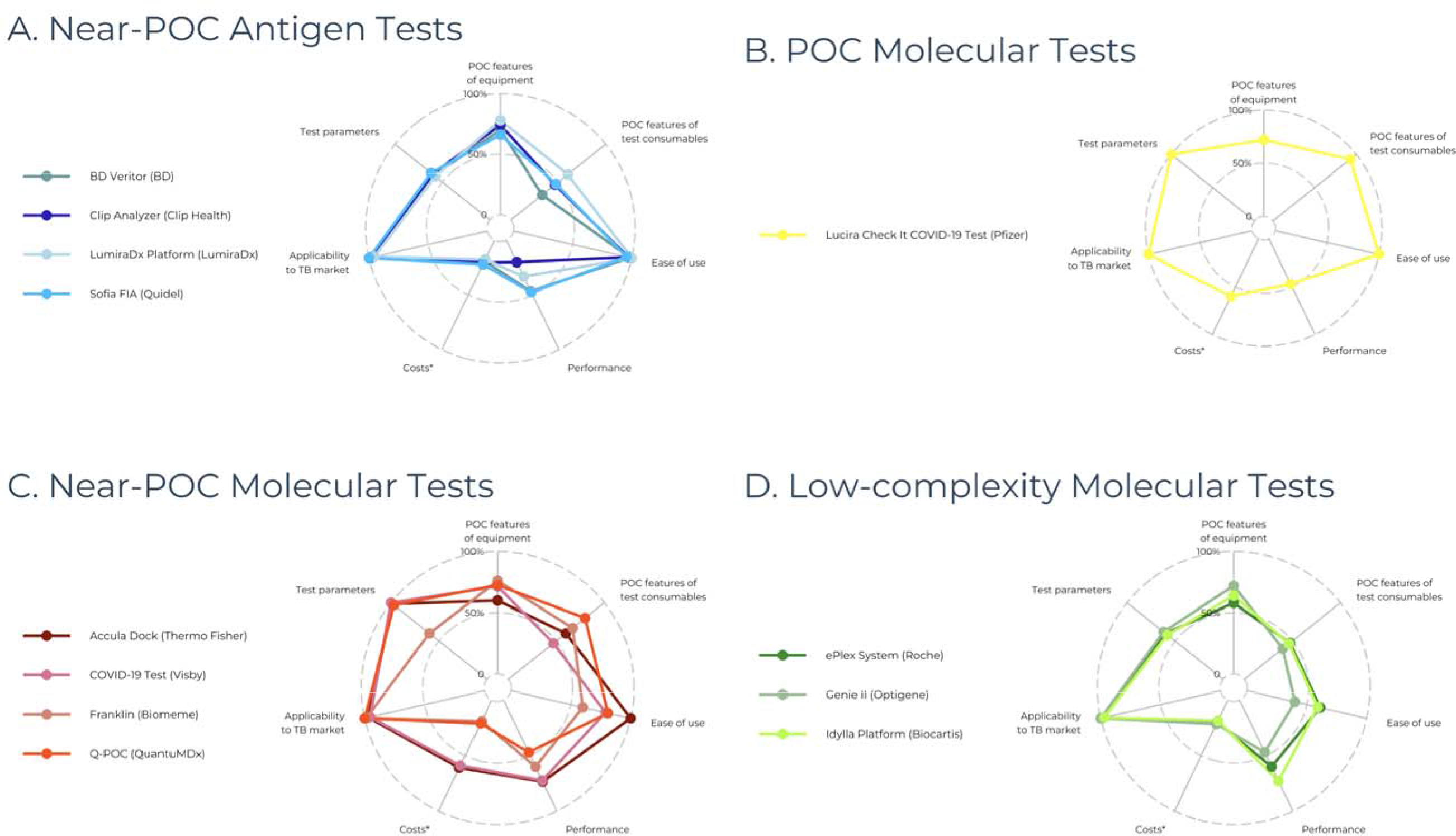
Performance of the three highest-scoring diagnostic devices across the seven scoring categories. Abbreviations: PoC = Point-Of-Care. Legend: *Costs (including capital costs and consumable costs) were not reported for most diagnostic tests, resulting in a score of 1/5 (20%).

#### 3.3.1 Near-POC Antigen Tests

We identified 17 tests that met the eligibility criteria. Six of those were reader-based LFTs, whereas 11 were automated immunoassays. Even though the Covid-19 Home Test (Ellume Health, Australia) is an LFT, we included it in this category since result interpretation requires a mobile phone. Table 2 summarizes the characteristics of the highest-scoring near-POC antigen test, the LumiraDx (LumiraDx, UK, recently acquired by Roche Diagnostics, Switzerland)[19], a multi-use, microfluidic immunofluorescence assay intended for the qualitative detection of antigens in non-sputum nasal swab (NS) and nasopharyngeal samples (NPS).

#### 3.3.2 POC Molecular Tests

We identified one true POC molecular test that requires two AA batteries to operate and is disposable: Lucira Check It COVID-19 Test (Pfizer, NY, USA). It can be used for the qualitative detection of RNA in nasal swab (NS) samples.

#### 3.3.2 Near-POC Molecular Tests

All 29 near-POC molecular tests included in the review were based on either PCR (n = 25 tests) or isothermal amplification (n = 5 tests). The vast majority of these tests are tabletop platforms, although we identified some smaller, handheld platforms like Cue Reader (Cue, CA, USA), DoctorVida Pocket Test (STAB Vida, Portugal) and Accula Dock (Thermo Fisher Scientific, MA, USA). We identified one disposable molecular test: Visby COVID-19 Test (Visby, CA, USA). Even though the test is disposable, because it requires stable power, we included it in this category.

#### 3.3.3 Low-Complexity Molecular Tests

As with the already mentioned molecular tests, the low-complexity molecular tests we identified were also based on either PCR (n = 7 tests) or isothermal amplification technology (n = 4 tests). The highest-scoring in this diagnostic category was Idylla^TM^ (Biocartis, Switzerland), a multi-use tabletop platform, weighing 18kg and running on standard electricity that is designed for the detection of RNA in NPS.

## 4 DISCUSSION

In this comprehensive scoping review, we identified 58 commercially available molecular and antigen tests for the diagnosis of SARS-CoV-2 at POC and assessed their applicability to TB. Our findings reveal a diverse array of diagnostic tests and instruments that hold potential in meeting the requirements of peripheral TB diagnostic testing.

### NEAR-POC ANTIGEN TESTS

The identified instrument-based near-POC antigen tests offer potential for meeting TPP sensitivity targets for TB antigen detection by employing signal-amplifying technologies, like readers paired with lateral flow assays and automated immunoassays utilizing sensitive detection methods, including fluorescence and electrochemical approaches. The front- runner, the LumiraDx, shows promising clinical performance with a low LoD of 2-56 PFU/mL in direct comparison with other tests[20]. However, reported diagnostic sensitivity of COVID- 19 assays included in the review varied widely, ranging from 37.5% to 90.0%, with limited data on LoD. This complicates the assessment of their potential to detect low-abundance TB antigens like lipoarabinomannan (LAM) in urine, where an LoD in the low pg/mL range is likely required so that the test can be used to detect TB in all patient groups [21]. To enable meaningful comparisons between assays, standardized LoD reporting is required[22]. Moreover, none of the identified platforms reported to use urine samples [23, 24]. As a result, successful application of identified platforms to TB will depend on optimized sample pre-treatment and concentration methods, specific anti-LAM antibodies, and sensitive readout approaches.

Overall, identified antigen tests excel in compact design, portability, and rapid turnaround- times, suited for decentralized settings. The LumiraDx is notable for its quick turnaround time, battery-operation, and multiple data export options, reducing reliance on Wi-Fi, though it only operates within limited temperature ranges[25]. Designing TB diagnostics must consider high temperatures and humidity in TB-endemic countries. Many identified tests also support multi-disease testing, challenging siloed testing programs, and facilitating differential diagnosis[26].

### POC AND NEAR-POC MOLECULAR TESTS

POC and near-POC molecular tests highlighted in this review present a variety of innovative assay technologies and platform features. These are designed to enhance user friendliness and testing capacity, incorporating features such as easy handling, self-testing options, rapid turnaround times, and the ability to detect multiple pathogens using multi-disease panels. The surge of new diagnostic tests has significantly boosted the scale up of SARS-CoV-2 diagnostic testing. However, adapting these molecular platforms for TB detection presents technical challenges due to Mycobacterium tuberculosis (Mtb)’s complex cell wall and low bacterial loads in clinical samples[6]. Firstly, obtaining non-sputum samples like tongue swabs, breath aerosols (XBA), and stool samples with high bacterial loads requires optimized collection to maximize bacterial capture[6, 27, 28]. Recent research suggests almost equal sensitivity of tongue swab PCR compared to sputum in symptomatic patients with “low” or higher sputum bacillary loads[29]. More research is needed for “very low” and “trace” sputum bacillary load cases. Most identified near-POC molecular tests support oral swabs, indicating adaptability to TB. Further, advances in aerosol collection using POC face masks in combination with cartridge-based NAATs make XBA a promising specimen, but sample processing needs streamlining[30, 31]. Additionally, stool samples’ acceptable sensitivity in children and simplified processing methods could enhance POC feasibility[32–34]. Despite being less sensitive than sputum samples, these less-invasive methods paired with molecular platforms suitable for use at POC could enhance diagnostic yield and patient acceptability and reduce overprescribing of empiric antibiotics[35, 36].

Secondly, while most identified molecular tests employ enzymatic or chemical lysis to release SARS-CoV-2 nucleic acids, mechanical lysis methods, like bead beating or sonication to break down Mtb’s lipid-rich cell wall are likely to be required [6]. Integration of these methods into a POC platform can be challenging. Alternatively, an accompanying POC device for mechanical lysis from swab samples could be envisioned, provided that the ease-of-use of the overall sample-to-result workflow is sustained [37, 38]. With tongue swabs, nucleic acid extraction may be skipped if cell lysis is efficient, simplifying the workflow[29]. Lastly, high-yield sample lysis must pair with sensitive molecular detection methods. For instance, the frontrunner candidate, Lucira Check It uses RT-LAMP, while Visby COVID-19 Test employs RT-PCR. Currently available isothermal amplification-based TB assays show high sensitivity but are limited in peripheral settings due to manual processes and outsourced DNA lysis and extraction[39, 40]. Integrating these assays with sensitive POC platforms, as identified here, could streamline testing.

Operational limitations of molecular platforms identified in this review may hinder their widespread adoption in peripheral settings. Some, like Visby COVID-19 Test and Accula Dock require a standard power source, affecting implementation in areas with unstable power supply[41]. Lucira Check It and Cue Reader address this with AA batteries or smartphone battery charge, but reliance on smartphones may still pose limitations. Further, most tests, including Lucira Check It and Visby COVID-19 Test, lack adequate data export options to reduce reliance on Wi-Fi connectivity. This raises concerns about manual result documentation, increasing the risk of human error and data loss, and hindering communities’ ability to fully leverage generated data[41–43]. Adapting to the needs of areas with limited internet and electronic medical record systems is essential to overcome these challenges[42]. Finally, many promising near-POC platforms have low daily sample throughput and limited multi-use capacity, hindering parallel sample analysis and potentially causing treatment delays. Among these platforms, Franklin^TM^ (Biomeme, PA, US) stands out, offering the detection of up to 27 targets in 9 samples per PCR run, including sexually transmitted-, mosquito-borne-, and respiratory pathogens. Multi-disease panels are crucial for integrated public health interventions and should be prioritized in the development of novel diagnostics[44]. Co-testing for TB, HIV, diabetes, and respiratory pathogens, in particular, could significantly benefit national health programs, guided by the WHO’s essential diagnostics list[45].

The at-home molecular tests suitable for self-testing, such as Visby COVID-19 Test, Lucira Check It, and Cue Reader show significant promise to meet TPP criteria. Their palm-sized format, minimal sample pre-processing, rapid result delivery, and minimal user training render them ideal for decentralized settings. To fully realize their potential, temperature and humidity ranges should be considered in TB-endemic settings, similar to the identified antigen tests. The recent shutdown of Cue Heath highlights the importance of expanding COVID-19 test platforms to other disease areas, such as TB, to capitalize on the rapidly growing TB diagnostic market[46].

### LOW-COMPLEXITY MOLECULAR TESTS

The GENIE® II (Optigene, UK), ranked second among the low-complexity platforms, holds potential to bridge gaps left by current WHO-recommended molecular systems[47]. It is battery-powered, operates at high temperatures (40 ℃), has a rapid 30-minute turnaround time for 16 samples, and accommodates USB data export. Additional LAMP primers can be designed to expand the diagnostic panel[48]. However, the instrument necessitates additional equipment for sample pre-treatment, limiting peripheral deployment, though simplifying sample preparation could improve this. Conversely, other highly ranked platforms, ePlex System (Roche Diagnostics, Switzerland) and Idylla^TM^, though not requiring sample pre-treatment, need continuous power and only operate at temperatures up to 30 ℃, with longer turnaround times (90 to 120 minutes) for 3 to 8 samples. Their multiplexing capacity and high throughput suit urban centers with substantial test volumes and laboratory infrastructure, similar to GeneXpert Dx[26].

### GENERAL FINDINGS

Overall, we observed a lack of transparency in the reporting of instrument and test costs, and where reported, they often exceeded WHO pricing recommendations. Currently, equitable access to WHO-recommended rapid molecular tests remains elusive in LMICs despite large-scale investments and price negotiations led by multiple stakeholders[26, 49, 50]. Addressing global affordability and accessibility requires diversified diagnostic manufacturing and minimized maintenance requirements[51]. However, most COVID-19 test manufacturers are in high-income countries, hindering global access[51]. Translation of identified tests to TB would require commitment from these companies to global health and global access terms. SD Biosensor’s (Suwon, South Korea) recent license agreement with the COVID-19 Technology Access Pool for its COVID-19 antigen test, under which nonexclusive sublicensees will receive the knowledge and materials to manufacture the technology could serve as a model for TB diagnostics[52]. Lastly, many identified tests lack independently reported clinical performance estimates, or discrepancies between developer- and independently reported values exist. This is in line with recent findings on overestimated developer-reported sensitivity estimates of rapid SARS-CoV-2 antigen tests [53].

This scoping review has multiple strengths. We conducted inclusive searches across various sources, including published studies, pre-prints, regulatory databases, and IVD manufacturers websites. This ensures a broad coverage of technologies from diverse developers, including start-ups, and large-scale IVD corporations. Data accuracy was ensured through rigorous screening conducted by two independent reviewers and cross-verification of charted data with developer-reported information. Lastly, objective scoring aligned with WHO TPPs was used to mitigate subjective reporting and to evaluate test characteristics of POC diagnostic devices against WHO standards.

Several limitations should be noted. First, due to the extensive dataset and time constraints, charted data were not cross-verified by a second reviewer, and early-stage non- commercialized platforms were excluded, potentially overlooking promising new technologies. Second, our search of IVD databases was confined to the openly accessible ones with English search functionality, resulting in a potential bias. Further, the scoring criteria based on Lehe et al’s scorecard require refinement, including weighing individual scores based on their relative importance to end-users in peripheral settings, and including additional TPP parameters. Also, the chosen scoring criteria were not specifically tailored to the various technology classes and test classifications. Some criteria are more applicable to certain technology classes than others. Lastly, data limitations, such as untransparent cost reporting and lack of clinical and analytical performance data may introduce bias in device scoring.

## 5 CONCLUSIONS

This scoping review has highlighted the potential for adapting SARS-CoV-2 POC diagnostic technologies for TB diagnosis, identifying 58 commercially available molecular and antigen tests that may meet WHO TPPs for peripheral settings. Despite the promising potential of several platforms, challenges remain, such as the need for optimized sample pre-treatment and considerations for deployment in resource-limited settings with varying environmental conditions. Additionally, addressing affordability and accessibility through diversified manufacturing and global health commitments is essential. Pricing structures must reflect the value across diverse settings, justifying costs with benefits like increased decentralization and enhanced clinical utility. Context-adapted diagnostic tests that integrate into local TB diagnostic algorithms and policies are preferred over one-size-fits-all solutions. This review serves as a foundational step toward leveraging COVID-19 diagnostic innovations to bridge the TB diagnostic gap, urging stakeholders to foster collaborations that translate these findings into impactful TB diagnostic solutions.

## 6 STATEMENTS AND DECLARATIONS

### Ethics and Dissemination

This scoping review did not require ethical approval because it does not involve individual patient data and uses sources that are in the public domain.

### Patient and Public Involvement

### No patients were involved in the study’s design, planning, or conception. Funding Statement

This systematic review is funded by the National Institutes of Health (NIH) (funding reference number U01AI152087; Rapid Research in Diagnostics Development for Tuberculosis Network).

### Author’s Contributions

S.Y. developed the scoping review protocol. L.H. and S.Y. collected, screened, and extracted data. L.H., S.Y., S.J. and R.D. reviewed and analyzed the data. L.H. S.Y. and S.J. drafted the manuscript. All co-authors provided critical editing and review. The final version was approved by all co-authors prior to publication.

### Author’s contact

Lydia Holtgrewe (Corresponding author), Heidelberg University Hospital, Division of Infectious Diseases and Tropical Medicine, Im Neuenheimer Feld 324, 69120 Heidelberg, Germany, +49(0) 6221- 56-35091

Lydia Holtgrewe: Sonal Jain

Ralitza Dekova: Tobias Broger: Chris Isaacs: Payam Nahid

Adithya Cattamanchi: Claudia Denkinger: Seda Yerlikaya

### Competing Interest Statement

Authors declare no financial conflict of interest. T.B. holds patents in the fields of lipoarabinomannan detection and aerosol collection, and is a shareholder of Avelo Ltd, a Swiss diagnostic company. C.I. is the founder and director of Connected Diagnostics Limited, a UK-based commercial entity that assists companies with the development of diagnostic devices. C.D. is a member of the Scientific Advisory Committee of Avelo Ltd.

## Data Availability

All data produced in the present study are available upon reasonable request to the authors

## Acknowledgments

The abstract was partially generated by ChatGPT (powered by OpenAI’s language model, GPT-3.5; http://openai.com). The editing was performed by the human authors.

## Notes

### Clinical Protocols

https://pubmed.ncbi.nlm.nih.gov/36754560/

