## Supplementary Materials for "Innovative COVID-19 Point-of-Care Diagnostics Suitable for Tuberculosis Diagnosis: A Scoping Review"

### Supplementary Methods

#### Supplementary Methods, Section 1. R code for the medrxivr package

#References: https://ropensci.org/blog/2020/10/20/searching-medrxivr-and-biorxiv-preprint-data/ ; https://docs.ropensci.org/medrxivr/

\# Set working directory

setwd("C:/Users/user/Documents/R/20220505 Rxiv Search")

\# Install and load required packages

install_if_missing <- function(package_name) {

if (!require(package_name, character.only = TRUE)) {

install.packages(package_name, dependencies = TRUE)

library(package_name, character.only = TRUE)}}

packages <- c("medrxivr", "dplyr", "ggplot2")

lapply(packages, install_if_missing)

\# Load required libraries

library(medrxivr)

library(dplyr)

library(ggplot2)

\# bioRxiv - Reproducible searching and exporting

\# Retrieve preprint data within a specified date range from bioRxiv

preprint_data <- mx_api_content(server = "biorxiv", to_date = "2022-11-23")

\# Define search criteria

criteria <- list(

condition = c("2019 nCoV", "2019nCoV", "2019 novel coronavirus", "COVID-19", "covid-19", "COVID19", "covid 19", "new coronavirus", "novel coronavirus", "novel corona virus", "sars cov 2", "SARS-CoV-2", "severe acute respiratory syndrome coronavirus 2"),

tech = c("molecular", "isothermal", "PCR", "polymerase chain reaction", "LAMP", "CRISPR", "immunoassay", "antigen"),

setting = c("point of care", "POC", "near patient", "rapid test", "bedside test", "laboratory-independent", "point-of-care", "POCT", "portable"),

usecase = c("diagnos", "detect"))

\# Run searches based on different criteria

results <- lapply(criteria, function(query) {

mx_search(data = preprint_data, query = list(query))})

\# Combine results

combined_results <- do.call(rbind, results)

\# Export the combined results

mx_export(combined_results)

write.table(combined_results, "bioRxiv_data_20221123.txt", sep = ",", row.names = FALSE)

write.csv(combined_results, file = "bioRxiv_data_20221123.csv", sep = ",", row.names = FALSE)

\# Download the PDF for each record

mx_download(combined_results)

\# medrxiv - Reproducible searching and exporting

\# Retrieve preprint data within a specified date range from medrxiv

preprint_data <- mx_api_content(server = "medrxiv", to_date = "2022-11-23")

\# Define search criteria

criteria <- list(

condition = c("2019 nCoV", "2019nCoV", "2019 novel coronavirus", "COVID-19", "covid-19", "COVID19", "covid 19", "new coronavirus", "novel coronavirus", "novel corona virus", "sars cov 2", "SARS-CoV-2", "severe acute respiratory syndrome coronavirus 2"),

tech = c("molecular", "isothermal", "PCR", "polymerase chain reaction", "LAMP", "CRISPR", "immunoassay", "antigen"),

setting = c("point of care", "POC", "near patient", "rapid test", "bedside test", "laboratory-independent", "point-of-care", "POCT", "portable"),

usecase = c("diagnos", "detect"))

\# Run searches based on different criteria

results <- lapply(criteria, function(query) {

mx_search(data = preprint_data, query = list(query))})

\# Combine results

combined_results <- do.call(rbind, results)

\# Export the combined results

mx_export(combined_results)

write.table(combined_results, "medrxiv_data_20221123.txt", sep = ",", row.names = FALSE)

write.csv(combined_results, file = "medrxiv_data_20221123.csv", sep = ",", row.names = FALSE)

\# Download the PDF for each record

mx_download(combined_results)

#### SUPPLEMENTARY TABLES

##### **Table S1**. PRISMA-ScR Checklist^[1]^

**Preferred Reporting Items for Systematic reviews and Meta-Analyses extension for Scoping Reviews (PRISMA-ScR) Checklist**

| **SECTION** | **ITEM** | **PRISMA-ScR CHECKLIST ITEM** | **REPORTED ON PAGE #** |
| --- | --- | --- | --- |
| **TITLE** | | | |
| Title | 1 | Identify the report as a scoping review. | Click here to enter text. |
| **ABSTRACT** | | | |
| Structured summary | 2 | Provide a structured summary that includes (as applicable): background, objectives, eligibility criteria, sources of evidence, charting methods, results, and conclusions that relate to the review questions and objectives. | Click here to enter text. |
| **INTRODUCTION** | | | |
| Rationale | 3 | Describe the rationale for the review in the context of what is already known. Explain why the review questions/objectives lend themselves to a scoping review approach. | Click here to enter text. |
| Objectives | 4 | Provide an explicit statement of the questions and objectives being addressed with reference to their key elements (e.g., population or participants, concepts, and context) or other relevant key elements used to conceptualize the review questions and/or objectives. | Click here to enter text. |
| **METHODS** | | | |
| Protocol and registration | 5 | Indicate whether a review protocol exists; state if and where it can be accessed (e.g., a Web address); and if available, provide registration information, including the registration number. | Click here to enter text. |
| Eligibility criteria | 6 | Specify characteristics of the sources of evidence used as eligibility criteria (e.g., years considered, language, and publication status), and provide a rationale. | Click here to enter text. |
| Information sources* | 7 | Describe all information sources in the search (e.g., databases with dates of coverage and contact with authors to identify additional sources), as well as the date the most recent search was executed. | Click here to enter text. |
| Search | 8 | Present the full electronic search strategy for at least 1 database, including any limits used, such that it could be repeated. | Click here to enter text. |
| Selection of sources of evidence† | 9 | State the process for selecting sources of evidence (i.e., screening and eligibility) included in the scoping review. | Click here to enter text. |
| Data charting process‡ | 10 | Describe the methods of charting data from the included sources of evidence (e.g., calibrated forms or forms that have been tested by the team before their use, and whether data charting was done independently or in duplicate) and any processes for obtaining and confirming data from investigators. | Click here to enter text. |
| Data items | 11 | List and define all variables for which data were sought and any assumptions and simplifications made. | Click here to enter text. |
| Critical appraisal of individual sources of evidence§ | 12 | If done, provide a rationale for conducting a critical appraisal of included sources of evidence; describe the methods used and how this information was used in any data synthesis (if appropriate). | Click here to enter text. |
| Synthesis of results | 13 | Describe the methods of handling and summarizing the data that were charted. | Click here to enter text. |
| **RESULTS** | | | |
| Selection of sources of evidence | 14 | Give numbers of sources of evidence screened, assessed for eligibility, and included in the review, with reasons for exclusions at each stage, ideally using a flow diagram. | Click here to enter text. |
| Characteristics of sources of evidence | 15 | For each source of evidence, present characteristics for which data were charted and provide the citations. | Click here to enter text. |
| Critical appraisal within sources of evidence | 16 | If done, present data on critical appraisal of included sources of evidence (see item 12). | Click here to enter text. |
| Results of individual sources of evidence | 17 | For each included source of evidence, present the relevant data that were charted that relate to the review questions and objectives. | Click here to enter text. |
| Synthesis of results | 18 | Summarize and/or present the charting results as they relate to the review questions and objectives. | Click here to enter text. |
| **DISCUSSION** | | | |
| Summary of evidence | 19 | Summarize the main results (including an overview of concepts, themes, and types of evidence available), link to the review questions and objectives, and consider the relevance to key groups. | Click here to enter text. |
| Limitations | 20 | Discuss the limitations of the scoping review process. | Click here to enter text. |
| Conclusions | 21 | Provide a general interpretation of the results with respect to the review questions and objectives, as well as potential implications and/or next steps. | Click here to enter text. |
| **FUNDING** | | | |
| Funding | 22 | Describe sources of funding for the included sources of evidence, as well as sources of funding for the scoping review. Describe the role of the funders of the scoping review. | Click here to enter text. |

JBI = Joanna Briggs Institute; PRISMA-ScR = Preferred Reporting Items for Systematic reviews and Meta-Analyses extension for Scoping Reviews.

* Where *sources of evidence* (see second footnote) are compiled from, such as bibliographic databases, social media platforms, and Web sites.

† A more inclusive/heterogeneous term used to account for the different types of evidence or data sources (e.g., quantitative and/or qualitative research, expert opinion, and policy documents) that may be eligible in a scoping review as opposed to only studies. This is not to be confused with *information sources* (see first footnote).

‡ The frameworks by Arksey and O’Malley (6) and Levac and colleagues (7) and the JBI guidance (4, 5) refer to the process of data extraction in a scoping review as data charting*.*

§ The process of systematically examining research evidence to assess its validity, results, and relevance before using it to inform a decision. This term is used for items 12 and 19 instead of "risk of bias" (which is more applicable to systematic reviews of interventions) to include and acknowledge the various sources of evidence that may be used in a scoping review (e.g., quantitative and/or qualitative research, expert opinion, and policy document).

##### **Table S2**. Data Charting Form

| **Section # Item #** | | | **Question type** | **Answer options** |
| --- | --- | --- | --- | --- |
| **General study information** | #1 Study ID  #2 DOI  #3 Title  #4 First author  #5 Publication year  #6 Publication type  #7 Research type | | #1 Short answer  #2 Short answer  #3 Short answer  #4 Short answer  #5 Drop-down  #6 Drop-down  #7 Drop-down | #1 NA  #2 NA  #3 NA  #4 NA  #5 2020, 2021, 2022  #6 Peer-reviewed, Pre-print  #7 Analytical research paper, Clinical research paper, Economic evaluation, Narrative review, Qualitative research paper, Systematic review, Unclear, Other |
| **Clinical performance** | #1 Study country  #2 Study design  #3 Study population  #4 Sample size  #5 Sensitivity (95% CI)  #7 Specificity (95% CI)  #8 Conflict of interest  #9 Financial support  #10 Developer among authors | | #1 Drop-down  #2 Drow-down  #3 Drow-down  #4 Short answer  #5 Short answer  #6 Short answer  #7 Short answer  #8 Drop-down  #9 Drop-down  #10 Drop-down | #1 Afghanistan (...) Zimbabwe, Multi-country, Not reported, Not relevant  #2 Case-control, Case report/series, Cross-sectional, RCT, Systematic review, Unclear, Other  #3 Adults, Children, Mixed, Not reported  #4 NA  #5 NA  #6 NA  #7 NA  #8 Yes, No, Unclear  #9 Yes, No, Unclear  #10 Yes, No, Unclar |
| **Test characteristics** | #1 Developer  #2 Country  #3 Product name  #4 Alternative product name  #5 Product type  #6 Product category  #7 Product description  #8 Limit of detection (copies/mL)  #9 Sample type  #10 Sample preparation  #11 Footprint (mm)  #12 Multi-use platform  #13 Throughout capacity  #14 Time-to-result (min)  #15 Hands-on time (min)  #16 Connectivity  #17 Max operating temperature (degree celsius)  #18 Max operating humidity (%)  #19 Shelf-life  #20 Power requirement  #21 Test price ($)  #22 Instrument price ($) | | #1 Short answer  #2 Drop-down  #3 Short answer  #4 Short answer  #5 Drop-down  #6 Short answer  #7 Short answer  #8 Short answer  #9 Short answer  #10 Drop-down  #11 Short-answer  #12 Drop-down  #13 Short answer  #14 Short answer  #15 Short answer  #16 Drop-down  #17 Drop-down  #18 Drop-down  #19 Drop-down  #20 Drop-down  #21 Drop-down  #22 Drop-down | #1 NA  #2 Afghanistan (...) Zimbabwe  #3 NA  #4 NA  #5 Antigen test, Molecular test  #6 NA  #7 NA  #8 NA  #9 NA  #10 No manual steps, 1-2 steps, >2 steps, Unclear  #11 NA  #12 Yes, No, Unclear  #13 NA  #14 NA  #15 NA  #16 Yes, No, Unclear  #17 25, 30, 40, 50, Unclear, Other  #18 70, 90, Unclear, Other  #19 12, 24, Unclear, Other  #20 Standard electricity, Solar-powered, Battery-powered, None, Unclear, Other  #21 <$1, $1-$2, $2-$4, $4-$10, >$10, Unclear, Other  #22 <$20, $20-$100, $100-$500, $500-$1000, $1000-$5000, >$5000, Unclear, not applicable (instrument-free), Other |

##### ***Abbreviations****: NA = Not applicable; CI = Confidence Interval.*

##### **Table S3**. Product Information Form

| **Item #** |  | **Question type** | **Answer options** |
| --- | --- | --- | --- |
| #1 Reference source  #2 Developer  #3 Developer website  #4 Business type  #5 Country  #6 Product name  #7 Product description  #8 Product type  #9 Product category  #10 Sample type  #11 Sample preparation  #12 Sensitivity (95% CI)  #13 Specificity (95% CI)  #14 Limit of detection (copies/mL)  #15 Test price ($)  #16 Instrument price ($)  #17 Footprint (mm)  #18 Multi-use platform  #19 Throughput capacity  #20 Time-to-result (min)  #21 Hands-on time (min)  #22 Connectivity  #23 Max operating temperature (degree celsius)  #24 Max operating humidity (%)  #25 Shelf-life  #26 Power requirements  #27 Technology readiness level (TRL)  #28 Regulatory status  #29 Potential use case  #30 Potential end user | | #1 Checkboxes  #2 Short answer  #3 Short answer  #4 Dropdown  #5 Drop-down  #6 Short answer  #7 Short answer  #8 Drop-down  #9 Checkboxes  #10 Checkboxes  #11 Drop-down  #12 Short answer  #13 Short answer  #14 Short answer  #15 Drop-down  #16 Drop-down  #17 Short answer  #18 Drop-down  #19 Short answer  #20 Short answer  #21 Short answer  #22 Drop-down  #23 Drop-down  #24 Drop-down  #25 Drop-down  #26 Drop-down  #27 Drop-down  #28 Drop-down  #29 Checkboxes  #30 Drop-down | #1 Developer website, FDA, EUDAMED, NMPA, MFDS, MDALL, CDSCO, FIND, Johns Jopkins, RADx  #2 NA  #3 NA  #4 Academia, Start-up, Not-for-profit, Micro company (<10), Small-sized company (10-49), Medium-sized company (50-250), Large-sized company (>250)  #5 Afghanistan (...) Zimbabwe  #6 NA  #7 NA  #8 Antigen test, Molecular test  #9 Automated immunoassay, CRISPR, High-throughput molecular test, Isothermal amplification, Rapid biosensor, Rapid molecular platform, Reader-based lateral flow assay, Vertical flow assay, Other  #10 Breath, Bronchoalveolar lavage, Feces, Mid-turbinate swab, Nasal swab, Nasopharyngeal sample, Oropharyngeal sample, Purified RNA, Saliva, Sputum, Whole blood, Other  #11 No manual steps, 1-2 steps, >2 steps, Unclear  #12 NA  #13 NA  #14 NA  #15 <$1, $1-$2, $2-$4, $4-$10, >$10, Unclear, Other  #16 <$20, $20-$100, $100-$500, $500-$1000, $1000-$5000, >$5000, Unclear, Not applicable (instrument-free), Other  #17 NA  #18 Yes, No, Unclear  #19 NA  #20 NA  #21 NA  #22 Yes, No, Unclear  #23 5, 30, 40, 50, Unclear, Other  #24 70, 90, Unclear, Other  #25 12, 24, Unclear, Other  #26 Standard electricity, Solar-powered, Battery-powered, None, Unclear, Other  #27 TRL1 (...) TRL9, Unclear  #28 CE-marked, Korea MFDS, US FDA EUA, US FDA 510k, WHO EUL, WHO-endorsed, Research use only, Under development, Unclear  #29 Detection, Triage, Drug susceptibility testing  #30 Trained laboratory technician, Healthcare workers trained to the level of auxiliary nurses, Healthcare workers with a minimum of training, Community or healthcare workers with a minimum of training, Self-testing, Other |

##### ***Abbreviations****: NA = Not applicable.*

##### **Table S4**. Scoring framework (adapted from *Lehe et al*)^[2]^

| **Scoring categories** | **Scoring criteria** | **Scoring variables** | **Definition of scoring criteria** | **Scoring thresholds** |
| --- | --- | --- | --- | --- |
| **POC features**  **of equipment** | Technical specifications | Instrument size | *Instrument size in cm.* | Disposable  =  5  Handheld  =  3  Tabletop  =  1 |
|  |  | Instrument weight | *Instrument weight in kg.* | Could be transported by hand (<5 kg)  =  5  Could be transported by vehicle (<15 kg) = 3  Could not be transported (>15 kg) = 1 |
|  |  | Power requirements | *Power source required to run the diagnostic device.* | Instrument-free = 5  Instrument with optional battery-powered operation = 4  Instrument with optional solar-powered operation = 3  Instrument running on standard electricity plus an uninterrupted power supply unit = 2  Instrument running on standard electricity only = 1 |
|  |  | Instrument-free | *Requirement of an instrument to run the diagnostic assay.* | Not required = 5  Required = 1 |
|  |  | Connectivity | *The diagnostic device can be connected to external devices, databases and/or cloud services via wifi or bluetooth connectivity. No manual data transfer via USB stick or other means is required.* | True = 5,  False = 1 |
|  | Data analysis | Integrated data analysis | *Data analysis and result display are integrated into the diagnostic device and do not require external devices.* | True = 5,  False = 1 |
|  |  | Integrated electronics and software | *The electronics and software required to process and display the test data are integrated into the diagnostic device.* | True = 5,  False = 1 |
|  | Testing capacity | Time-to-result | *Time passed between sample collection and result display in minutes/hours. When a range was reported, the upper limit was used as a reference value for scoring.* | <15min = 5  <30 min = 4  <1 hour = 3  <2 hours = 2  >= 2 hours = 1 |
|  |  | Hands-on-time | *Time passed between sample collection and automated sample processing by the diagnostic device in minutes/hours. When a range was reported, the upper limit was used as a reference value for scoring.* | <1 min = 5  <5 min = 4  <10 min = 3  >10 min = 1 |
|  |  | Throughput capacity | *Number of samples that can be processed by the diagnostic device in a single run.* | ≥ 2 tests per run = 5  1 test per run = 1 |
| **POC features of test consumables** | Operating conditions | Operating Temperature | *The maximum allowable temperature (°C) of the local ambient environment at which the diagnostic device can be operated.* | Up to 50°C = 5  Up to 40°C = 4  Up to 30°C = 3  Up to 25°C = 2  <25°C = 1 |
|  |  | Operating Humidity | *The maximum allowable humidity (%) of the local ambient environment at which the diagnostic device can be operated.* | Up to 90% humidity = 5  Up to 70% humidity = 3  <70% humidity = 1 |
|  | Storage conditions | Shelf life | *The length of time (months) during which the diagnostic assay can be stored without becoming unfit for diagnostic testing, i.e. without compromising on accuracy.* | >12 Months  =  5  6–12 Months  =  3  <6 Months  =  1 |
| **Ease of use** | End user requirements | Potential end user | *Level of professional training required of the diagnostic device's potential end user:*  *1. Community or lay health worker without technical skills includes diagnostic devices for which no special technical skills are required, including those suitable for self-testing*  *2. Healthcare workers with basic technical skills include diagnostic devices that require basic technical skills. Devices in this category do require pre-processing of the sample that can be performed without professional laboratory equipment.*  *3. Healthcare workers with more advanced technical skills include diagnostic devices that require more advanced technical skills. Devices falling into this category require extensive sample pre-processing, including laboratory equipment, such as exact pipetting and precise volume transfer.* | Community/lay health worker without technical skills = 5  Healthcare worker with basic technical skills = 3  Healthcare worker with more advanced technical skills = 1 |
|  |  | Number of Manual Sample Processing Steps | *The number of sample processing steps that must be performed manually between sample collection and automated sample processing by the diagnostic device. This excludes the sample collection itself, but includes steps such as pipetting, addition of buffers, or vortexing.* | No manual steps = 5  1-2 steps = 3  > 2 steps = 1 |
| **Performance** | Analytical and clinical performance (COVID-19) | Limit of detection | *The smallest concentration of analyte that can be detected by the diagnostic assay. For the purpose of scoring, the lowest reported LoD (copies/mL) was used. Only study-reported estimates were considered.* | ≤ 100 copies/mL = 5  ≤ 500 copies/mL = 3  > 500 copies/mL = 1 |
|  |  | Clinical sensitivity | *The probability of a positive test results in truly positive individuals. For scoring purposes, an estimate extracted from systematic reviews was used. If only estimates from cross-sectional studies or case-control studies were available, estimates from cross-sectional studies were preferred. If multiple estimates were available, the estimate from the study with the largest sample size was selected. Only study-reported estimates were considered.* | > 98 % = 5  > 95 % = 3  < 95 % = 1 |
|  |  | Clinical specificity | *The probability of a negative test result in truly negative individuals. For scoring purposes, an estimate extracted from systematic reviews was used. If only estimates from cross-sectional studies or case-control studies were available, estimates from cross-sectional studies were preferred. If multiple estimates were available, the estimate from the study with the largest sample size was selected. Only study-reported estimates were considered.* | > 98 % = 5  > 95 % = 3  < 95 % = 1 |
| **Cost** | Upfront and user costs | Capital cost of equipment | *Fixed, one-time upfront costs that are incurred when purchasing the diagnostic device. Instrument-free diagnostics received a score of 5.* | <$1,000  =  5  $1,000–5,000  =  3  >$5,000  =  1 |
|  |  | Consumable cost | *Recurring costs to run a single diagnostic test.* | <$2 = 5  $2–10 per test  =  3  >$10 per test  = 1 |
| **Applicability of platform to TB market** | Multi-use | Applicability of platform to other indications | *The ability to run diagnostic assays for different indications on the diagnostic device. Such assays can for example be Influenza or CMV assays.* | True = 5  False = 1 |
| **Others** | Test parameters | Number of test  parameters  available | *Number of scoring variables available for the diagnostic device compared to the average number of scoring variables available within the test category.* | >18 parameters available =  5  12-18 parameters available  =  3  <12 parameters available  =  1 |

##### ***Abbreviations****: PoC = Point-of-Care; NA = Not applicable.*

##### Table S5. Near-POC Antigen Tests.

[Please see separately attached excel file]

##### Table S6. POC Molecular Tests.

[Please see separately attached excel file]

##### Table S7. Near-POC Molecular Tests.

[Please see separately attached excel file]

##### Table S8. Low-complexity Molecular Tests.

[Please see separately attached excel file]
